# A novel approach to determine the micromotion of screw-plate interfaces of a locking-type plating system under axial cyclic loading via synchronized digital image correlation

**DOI:** 10.1101/2025.04.11.25325637

**Authors:** Christian Halbauer, Andreas Paech, Felix Capanni

## Abstract

**Objectives:** A previously published biomechanical study of axial cyclic testing on a locking-type plating system presented several failure types of the screw-plate interfaces (SPI). It was assumed that increasing micromotions of SPIs result consequently in initial failure of SPIs during cyclic loading. In conclusion, measurements of SPIs via digital image correlation (DIC) were suggested to detect potential micromotions.

**Methods:** DIC measurements were performed using the ARAMIS Adjustable 3D measurement system to track the surface of the screw-head and bone plate during cyclic testing to determine potential micromotions. The micromotion is thereby described as the change in distance of the screw-head center to a reference point on the bone plate, providing information about the quality of interlocking of the related SPI during cyclic loading. Micromotion analysis was performed on the fracture-adjacent SPIs of the implant system, that were considered as the most relevant interfaces.

**Results:** Micromotion could be detected for both facture-adjacent SPIs in all test samples with increasing magnitude during cyclic testing, resulting in micromotions up to 587 µm for the proximal and 321 µm for the distal fracture-adjacent SPI after 50k load cycles. The strongest increase in micromotion could thereby be detected within early stages of cyclic loading.

**Conclusions:** The approach to determine micromotions of SPIs during axial cyclic loading was successful, indicating a reduction of the SPI’ s quality of interlocking as micromotions increase during cyclic loading of the tested locking-type implant system.

## Introduction

Osteosynthesis with locking plates is a common method to treat fractured bones in surgery. In the year 2023 alone, plating osteosynthesis via locking-type implant systems was performed 17,576 times for fracture treatments of long bones in Germany [1]. Thereby, the construct of a locking-type bone plate and several locking-type screws bridges the fracture, aligns the fractured bone in the anatomically correct position and ensures stability during rehabilitation and fracture healing [2]. Therefore, the implant system needs to withstand postoperative physiological loads within sporadic and repetitive load scenarios, as the acting loads need to be transferred through the implant system between fracture sides [3]. To ensure safety requirements, implant system testing is essential to evaluate the biomechanical performance under static and cyclic load conditions within physiological boundaries [3–15].

The locking-type connection of the bone plate and the screw – the screw-plate interface (SPI) – is therefore a crucial interface of the implant system, as failure of single or multiple SPIs reduces the construct stiffness, leading to a reduction in performance that might result in total failure of the implant system.

In a previously published study of the authorship [12], a biomechanical investigation regarding the impact of varying load and support conditions in implant system testing was conducted on the locking-type plating system tifix® femoral wave 8-hole by litos/ (litos/ GmbH, Ahrensburg, Germany) for femoral shaft fractures under physiological boundaries. Biomechanical testing resulted in multiple failure types of SPIs that accumulated into total failure of the implant system in a series of test samples under continuous cyclic loading. It was observed that the initial failure of SPIs originated by a so-called pop-up of the screw head of the related SPI, as the screw head’ s thread lost connection to the bone plate and elevated upwards due to the overall bending of the test sample. It was further postulated that this failure type might be linked to potential micromotions of the screw head with increasing characteristics during cyclic loading, resulting consequently in the initial pop-up failure of SPIs. To proof this theory, measurements via digital image correlation (DIC) were suggested to track the relative movement of the screw head of SPIs during axial cyclic loading, as a determination of the quality of interlocking and its change during testing. And to the authorship’ s knowledge, there is no study available in literature covering this specific topic.

In that context, this study presents a novel approach to determine the micromotion of SPIs of a locking-type plating system under axial cyclic loading via synchronized DIC. The presented study is linked to the preceding study [12], as the DIC measurements were performed synchronously to biomechanical testing as a proof of concept. The study covered the following key research questions: (1) Is it possible to detect micromotion of SPIs during cyclic loading using a DIC-based 3D measurement system?, (2) Does the magnitude of micromotions change during cyclic loading? And (3) Are micromotion measurements suited to be used as an indicator regarding the quality of interlocking of SPIs?

## Materials and Methods

The following sections provide a detailed description of the configuration of the test setup, the approach of detecting micromotions of SPIs via DIC, the performed test procedure and data acquisition of the presented study, that are based on a previously published study on axial IST [12].

### Test setup configuration

An axial implant system test was performed under physiological load and support conditions, as were presented in a previously published study on an investigation regarding the impact of different physiological boundary conditions on axial implant system testing [12]. The full test setup is displayed in Figure 1.

**Figure 1:**
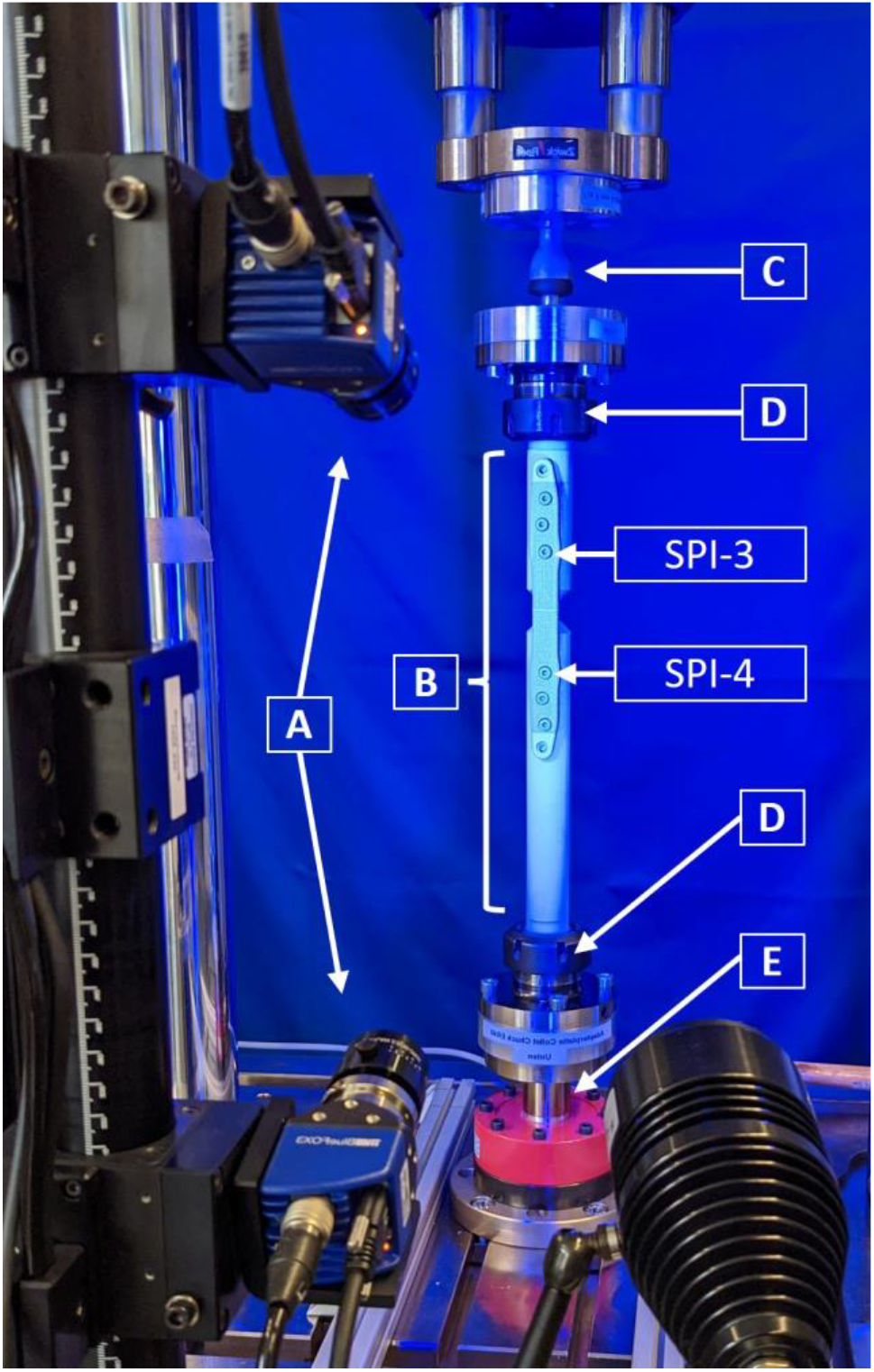
Test setup of the axial implant system test with synchronized digital image correlation using the ZEISS ARAMIS Adjustable 3D measuring system [A], tracking the motion of the implant system [B] during axial cyclic loading, applied trough a spherical load support at the proximal top [C] while mounted between two collet chucks [D] to a the fixed support at the distal end at the load cell [E], to measure the micromotions of the fracture-adjacent screw-plate interfaces – SPI-3 and SPI-4.

A custom made straight and unbend version of the locking-type plating system tifix® femoral wave 8hole by litos/ (litos/ GmbH, Ahrensburg, Germany) for femoral shaft fractures was tested under cyclic loading. Therefore, the implant system was mounted with a bridge-plating offset of 2 mm onto two cylindrical rods of Polyoxymethylene (POM) with a diameter of 30 mm. The test samples featured a 10 mm gap between the POM rods at the center of the implant system to demonstrate an AO32A diaphyseal fracture. The test samples were mounted via two collet chucks ER40 to an electrodynamic testing machine LTM-5 (ZwickRoell GmbH & Co. KG, Ulm, Germany), equipped with a 5 kN load cell. The load support at the proximal end of the test sample featured a ball-joint THK RBI 10BD, allowing free axial rotation and angular rotation up to 10 degrees perpendicular to the axial axis. The distal end of the test sample featured a fixed support, as the collet chuck was mounted directly to the load cell.

The 3D measuring system ARAMIS Adjustable by ZEISS (Carl Zeiss GOM Metrology GmbH, Braunschweig, Germany) was used to measure the micromotion of the fracture-adjacent SPIs (SPI-3 & SPI-4), as literature indicates that SPIs closest to the fracture bear the most load for similar test setup configurations [16–20] and failure of various interfaces of this implant system was repeatably detected in the preceding study [12]. The measuring system configuration featured two 2.3 Megapixel cameras with 35 mm lenses and the ARAMIS controller for synchronized recording. Stereo images were recorded at a sampling rate of 60 Hz. Calibration was performed on a measuring volume of 170 mm height, 110 mm width and 90 mm depth, resulting in a calibration deviation of 0.010 pixel and an accuracy of μm. The ARAMIS pattern printer JetStamp 1025 was used to ensure consistent quality of the spray pattern on all test samples.

Further information on the test setup configuration is available in the previously published study of biomechanical investigation on which this study is based on [12].

### Micromotion via DIC

In general, the 3D measuring system is capable of tracking pixel-facets over time in 3D space within the calibrated measuring volume via DIC. The center of each pixel-facet is expressed as a facet-point with coordinates in relation to the global coordinate system.

The principle of micromotion detection of an SPI is based on the relative movement of the center of the screw head in relation to a reference facet-point on the bone plate surface close to the corresponding interface. The change in distance between the center of the screw head and the reference point on the bone plate describes the quantity of micromotion in micrometer (µm). The principle is further explained in relation to the schematic visualization of Figure 2. To determine the center of a screw head (yellow dot), four facet-points (P1-P4) are defined on opposite corners of the hexagonal countersink of the screw head. The center of the screw head could be expressed as the crossing point of the vectors P1P3 (dashed black vector) and P4P2 (dashed blue vector). However, the two vectors are typically skewed to one another in 3D space and possess no intersection. Therefore, the center of the screw head (yellow dot) was defined as the center of the closest distance of the two skew vectors P1P3 and P4P2 in 3D space, as visualized in Figure 2.

**Figure 2:**
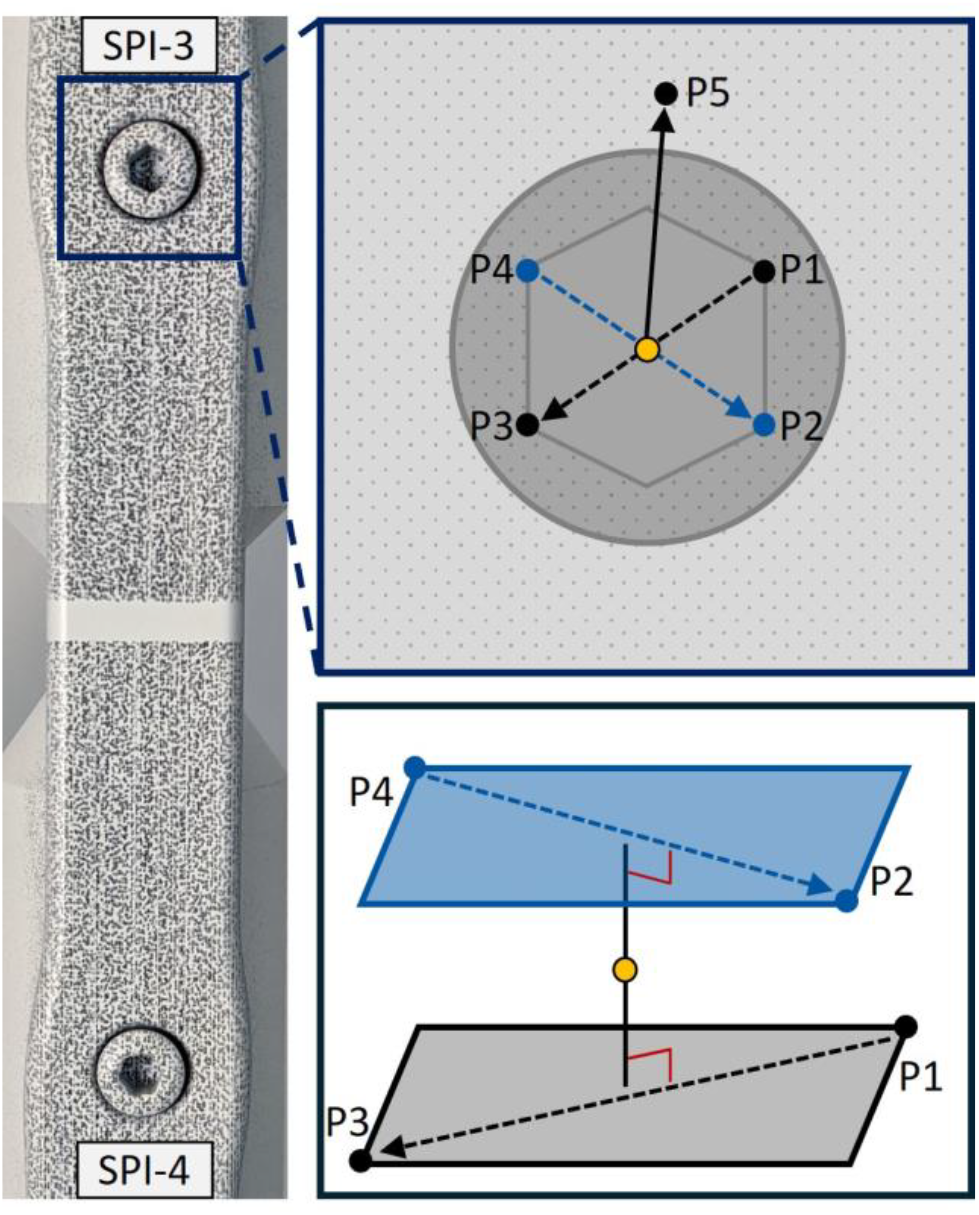
Schematic visualization of the approach to measure the micromotion of a screw-plate interface (SPI) via digital image correlation, as four points of the screw head’ s hexagonal countersink (P1-P4) and a fifth point (P5) on the bone plate surface are defined and tracked in 3D space, whereas the center of the screw head (yellow dot) is calculated as the center of the smallest distance between to two skew vectors P1P3 (dashed black vector) and P4P2 (dashed blue vector) in 3D space, defining the micromotion of the SPI as the change in length of the vector between the center of the screw head and the point P5 (black vector).

A fifth facet-point (P5) is defined on the bone plate surface close to the corresponding SPI to draw the resultant vector for micromotion detection (black vector) between P5 and the center of the screw head. In case of a change in the quality of interlocking of the screw head to the bone plate, the SPI’ s micromotion is described by the change in length of the resultant vector.

It is important to point out that strain effects of individual components may impact the micromotion measurements, as the bending related elongation of the bone plate could increase the micromotion vector’ s length. For that reason, the facet-point P5 was placed in direction of its neighboring screw, as it was hypothesized that the elongation of the bone plate in-between the three proximal and distal screws is negligible small. To check the hypothesis, strain analysis via DIC was performed for P5 of SPI-3 and SPI-4 for each test sample during axial IST, resulting in a negligible error in micromotion measurement as maximal local strains were smaller than 0.3%.

### Test procedure

Testing procedure was defined according to the previously published study of axial IST, on which this study is based on [12].

Six test samples (n = 6) were tested under sinusoidal cyclic axial loading to a physiological load limit of F_max_ = 1670 N at 2 Hz for 500k load cycles with a load ration of R = 0.1. Physiological loading is considered in terms of 90% of the average axial load maxima at the hip joint of a patient of 80 kg body weight during walking [12, 21–24].

Micromotion analysis via DIC was conducted for the range of the first 50k load cycles, as this range was identified as the load cycle range of initial failure of SPIs [12]. Within this range of load cycles, six measurements with the ARAMIS Adjustable 3D measurement system were recorded synchronously to the axial loading of each sample. Each recording tracked the test sample at a sampling rate of 60 Hz for 20 seconds, allowing micromotion analysis via DIC of 40 load cycles for each recording. The first recording tracked the initial 40 load cycles of testing. The other five recordings started at 10k, 20k, 40k and 50k load cycles to measure the progress of micromotion in comparison to the initial micromotion of the first recording. Micromotion analysis was conducted solely on the fracture-adjacent interfaces SPI-3 and SPI-4, as it was to be expected that the SPIs closest to the fracture bear the most load [16–20]. The fracture-adjacent proximal screw was defined as SPI-3 and the fracture-adjacent distal screw was defined as SPI-4, as seen in Figure 1 and Figure 2.

Synchronization between measuring systems was achieved by sending a 5V high-low trigger signal from the input/output card of the testing machine to the ARAMIS Controller during testing at the pre-defined load cycles of recording (1^st^, 10k, 20k, 30k 40k and 50k load cycle), starting an automated measurement procedure of ARAMIS. Before each recording, the ARAMIS system was set on hold, awaiting the trigger signal to start recording.

### Data acquisition

The software ZEISS INSPECT Correlate Pro (version 2025.1.0.1985) was used for the pre- and postprocessing of measurements and DIC analyses. Coordinates of facet-points were exported and further analyzed in MATLAB® R2024a (MathWorks®, Natick, MA, USA) to calculate the screw head center and the resultant micromotion.

## Results

Testing procedure with synchronized measuring systems was successfully performed throughout testing. Micromotion was detected for both interfaces SPI-3 and SPI-4 in all six test samples with increasing motion across recordings. In general, major changes in micromotion were detected in the first recording and after 10k load cycles in the second recording, as further changes in micromotion were comparably small in reference to the previous recording. As the resultant micromotion is expressed as positive and negative values depending on the direction of motion of the screw head center in relation to the reference facet-point P5, results of micromotion measurements are presented in Figure 3 as rational numbers. Yet, in order to describe the relative change in micromotion across recordings, data analysis was performed on the absolute values of the detected micromotions.

**Figure 3:**
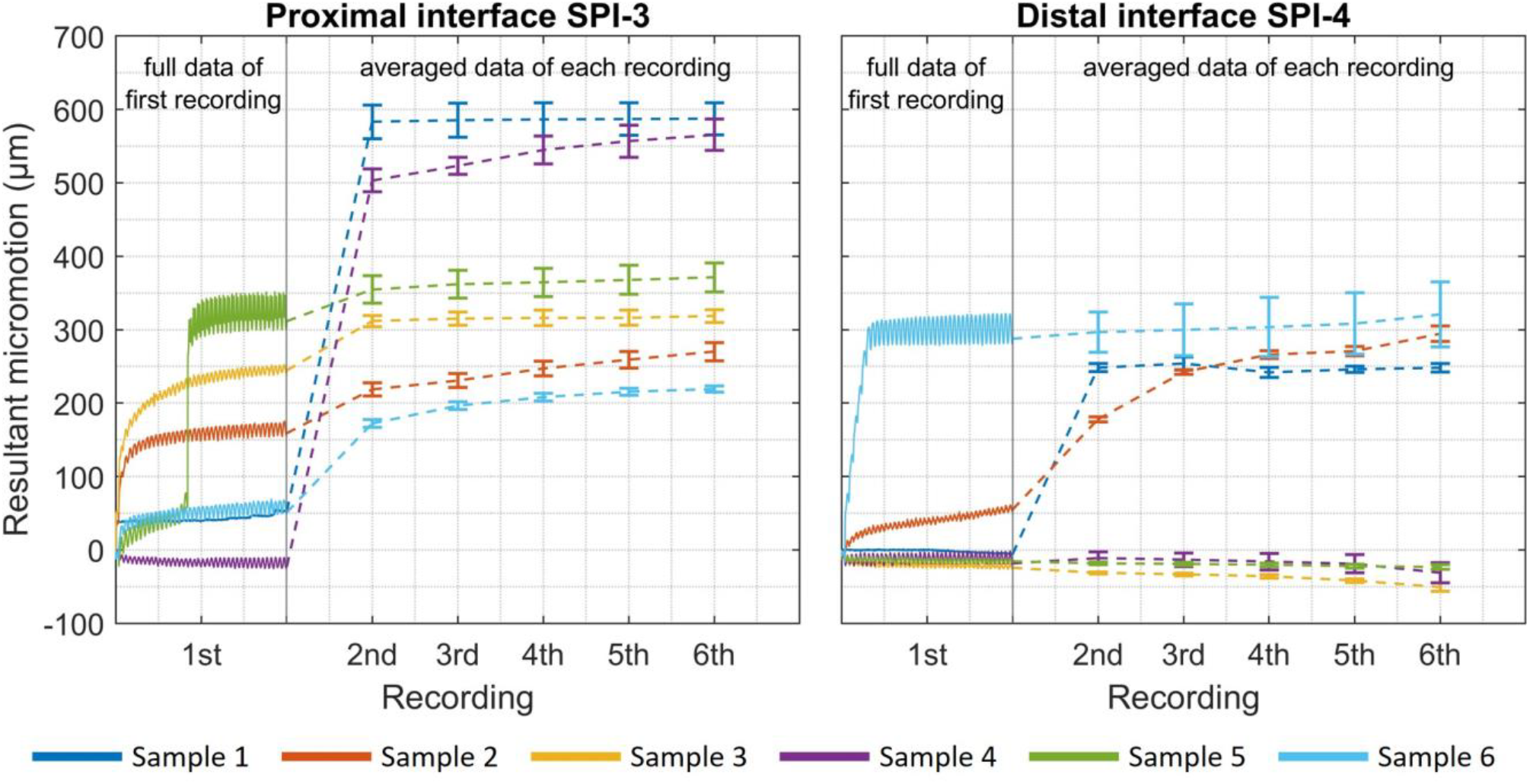
Resultant micromotion of the fracture-adjacent screw-plate interfaces SPI-3 and SPI-4, displayed as the full measurement data of the resultant micromotion during the first recording (initial 40 load cycles) and the averaged measurement data of the resultant micromotion of the second to the sixth recording, presenting the micromotion after intervalls of 10k load cycles in terms of the respective average value and the range of micromotion as error-bars.

In terms of the fracture-adjacent proximal SPI-3, an average micromotion of 144 μm was determined at the end of the first recording, showing a strong increase in micromotion in 5 of 6 test samples and reaching a micromotion maximum of 321 µm within the initial 40 load cycles for Sample 5. In terms of SPI-4, an average micromotion of 69 µm was determined at the end of the first recording, where only 2 of 6 test samples showed a strong increase, yet reaching a micromotion maximum of 302 µm within the initial 40 load cycles for Sample 6.

After 10k load cycles, the second recording revealed an average rise in micromotion of 219 µm in terms of SPI-3, and an average rise in micromotion of 66 µm for SPI-4. Thereby, a micromotion maximum was detected for SPI-3 in Sample 1 with 583 µm, and for SPI-4 in Sample 6 reaching 297 µm.

Further changes in micromotion of SPI-3 were comparably small yet steadily increasing across second to sixth recording, resulting in an average rise in micromotion of 8 µm per 10k load cycles in the range from 10k – 50k load cycles. In case of SPI-4, a similar characteristic was present, resulting in an average rise in micromotion of 9 µm per 10k load cycles.

The sixth recording resulted in an average micromotion after 50k load cycles of 389 µm for SPI-3, and 126 µm for SPI-4. However, 3 of the 6 test samples showed in terms of SPI-4 micromotions below 50 µm after 50k load cycles, whereas the other test samples reached micromotions above 240 µm.

## Discussion

The approach to determine micromotion of SPIs of a locking-type plating system under axial cyclic loading via synchronized digital image correlation was successful. The 3D measuring system ARAMIS Adjustable was capable of tracking the patterned surface of the implant system during axial cyclic loading and provided the global coordinates of each facet-point for determining the micromotion of the fracture-adjacent SPIs.

Measurement data analysis revealed micromotion in both interfaces SPI-3 and SPI-4 with similar characteristics, resulting in an average micromotion of 389 µm for SPI-3 and 126 µm for SPI-4 after 50k load cycles. In general, three major outcomes could be concluded that will be discussed in the following paragraphs.

First, micromotion could be detected within early stages of cyclic loading, as both SPIs showed micromotion within the initial 40 load cycles of testing in the first recording. Especially SPI-3 showed a strong increase in micromotion in 5 of the 6 test samples within the first recording. In terms of SPI-4, only 2 of the 6 test samples showed a strong increase in micromotion within the initial 40 load cycles.

Second, only minor changes in micromotion were detected across recordings after 10k load cycles, as an average rise in micromotion of 8 µm per 10k load cycles in terms of SPI-3, and 9 µm per 10k load cycles in terms of SPI-4, were detected. However, the recordings represent only a fraction of the resulting micromotion during cyclic loading. The instant or range of the switch from major to minor changes in micromotion was not captured, as each recording tracked intervals of 40 load cycles. It is reasonable to assume that this instant or range of load cycles is reached during early cyclic loadings and not until 10k load cycles, as the strong change in micromotion within the initial 40 load cycles indicates. It is therefore recommended to extend initial DIC measurements to determine a potential biphasic behavior of rising micromotion, describing the initial strong increase and the switch to small but steady increase in micromotion.

Third, SPI-3 experienced strong micromotions throughout all test samples, whereas SPI-4 showed similar micromotion quantities in only 3 of the 6 test samples, indicating a disequilibrium in resulting loads acting on the implant system, although being geometrically symmetrical. The disequilibrium can be explained by the different boundary conditions at the proximal and distal end of the test sample and their impact on the bending stiffness. In contrast to the fixed support at the distal end, the ball joint of the load support at the proximal end of the test sample allows free angular rotation, resulting in an overall smaller bending stiffness of the construct proximal of the fracture, compared to the construct distal of fracture, as the fracture is in the center of the test samples working length. Concludingly, due to the overall bending effect of the test sample, the proximal construct experienced larger deformations, resulting in higher loads at the interface SPI-3.

The findings of this study are linked to several limitations that need to be considered for evaluation. Despite the derivation of plausible physiological loading conditions and load limits, as was described in detail in the preceding study [12], the applied axial loading represents the main load component only. Therefore, a more complex load condition of 2D or 3D physiological loading might result in different micromotions of the two SPIs in magnitude and characteristic.

Another limitation is the restriction of the presented findings towards the tested implant system and test setup configuration. The tested implant system featured a multidirectional screw system, in which the thread at the screw head cuts into a narrow lip in the screw hole of the bone plate, creating an angle-stable connection. However, the connection is formed with little interacting material, that was considered as the weak spot of the implant system in the previous study [12], as multiple SPIs failed in testing. Therefore, the detected characteristic and magnitudes of micromotion accounts solely to that type of multidirectional fixation of this specific implant system. Other well established fixation systems feature unidirectional SPIs, such as the Locking Compression Plate System (LCP™ System) by DePuy Synthes (Johnson & Johnson, USA), in which the screw head’ s thread ties positively to the bone plate’ s thread, that might result in different magnitudes of micromotion when cyclically loaded. It is assumed that such unidirectional interfaces experience smaller micromotion under equal load condition of this study, as the interacting material of the components is larger, and loads are distributed more evenly. Yet, failure of unidirectional SPIs in form of pop-up failures during biomechanical testing can be found in literature [25]. It is also reasonable that different applications, such as plating osteosynthesis of the humerus and the femur, feature different magnitudes of micromotion, as dimensions of implant systems and physiological load conditions vary between applications. In that context, the determined micromotion of this study cannot be evaluated in terms of the magnitude and in relation to a critical threshold. Therefore, further biomechanical studies are needed to investigate micromotions of SPIs of different established plating systems with an application-wise scope to evaluate the quality of interlocking and critical magnitudes of micromotion.

In the previous study [12], DIC was considered as a promising approach to determine potential micromotions of SPIs to investigate the quality of interlocking of a locking-type screw head to the bone plate. By the findings of the presented study, the DIC approach is considered as a valid option to determine the quality of interlocking, whereas a rising micromotion of SPIs under cyclic loading might indicate the potential of failure of SPIs. As a major finding of the biomechanical investigation in the preceding study [12], total failure of the implant system was observed as an accumulation of different forms of failures of the SPIs and initial failure was described as a pop-up of the screw head. Considering the results of micromotion from this study, it is plausible to assume that the initial pop-up failure is the result of increasing micromotions, as the multidirectional connection between the screw head and bone plate fails due to slowly increasing plastic deformation in the SPI. Consequently, it should be concluded, the less micromotion an SPI experiences during cyclic loading, the better the quality of interlocking.

On the contrary, a controlled micromotion of SPIs might offer the opportunity to increase the interfragmentary movement of the fracture zone to promote fracture healing [26–30]. Innovative designs and prototypes of implant systems with the feature of controlled micromotion, as were presented over the past years to the biomechanical society [29, 31–33], could profit from the DIC approach of this study to evaluate the characteristics of micromotion and the impact on interfragmentary movements.

## Data Availability

All data produced in the present work are contained in the manuscript

## Acknowledgements

The authors thank the whole research team and supporters for its teamwork.

## Research funding

The research project was funded by the Federal Ministry for Economic Affairs and Energy and by the Central Innovation Program for small and medium sized enterprises of Germany.

